# Self-applied ELCSA is valid for rapid tracking of household food insecurity among pregnant women during the COVID-19 pandemic

**DOI:** 10.1101/2020.09.22.20199380

**Authors:** Thilini C. Agampodi, Amber Hromi-Fiedler, Suneth B. Agampodi, Gayani S. Amarasinghe, Nuwan D. wickramasinghe, Imasha U. Jayasinghe, Ayesh U. Hettiarachchi, Rafael Perez-Escamilla

## Abstract

**Background:** Rapid household food insecurity (HFI) tracking has been identified as a priority in the context of the COVID-19 pandemic and its aftermath. We report the validation of the Latin American and Caribbean Food Security Scale (*Escala Latinoamericana y Caribena de Seguridad Alimentaria -* ELCSA) among pregnant women in Sri Lanka.

**Methods:** The adult eight-items of the English version of ELCSA was translated into Sinhala and Tamil. Cognitive testing (on ten pregnant women and five local experts) and psychometric validation of the self-administered HFI tool was conducted among pregnant women (n=269) attending the Rajarata Pregnancy Cohort (RaPCo) special clinics in Anuradhapura in February 2020. We assessed psychometric properties and fit using a one parameter logistic model (Rasch analysis) using STATA version 14 and WINSTEP software version 4.3.4. Concurrent validity was tested using psychological distress.

**Results:** The scale was internally consistent (Cronbach’s alpha = 0.79), had a good model fit (Rasch infit statistic range: 0.85 to 1.07). Item 8 (‘did not eat for the whole day’) was removed from the model fit analysis as it was not affirmed by anyone. Item severity scores ranged from -2.15 for ‘not eating a diverse diet’ to 4.43 for ‘not eating during the whole day’. Concurrent validity between HFI and psychological distress was confirmed (r=0.15, p<0.05).

**Conclusions:** The self-applied version of ELCSA-pregnancy in Sri Lanka (ELCSA-P-SL) is a valid and feasible tool to track HFI among pregnant women in similar contexts during the COVID-19 pandemic, where social distancing is a major concern and its aftermath.

## 1.0 Introduction

In high income countries (HICs) as well as low and middle income countries (LMICs) household food insecurity (HFI) has been associated with non-communicable diseases in pregnancy such as gestational diabetes mellites, pregnancy induced hypertension and obesity (1,2). A recent systematic review in South Asia indicate that HFI is a determinant of intra household food allocation with women acting as a buffer and being more sensitive to deprivation of food (3). In LMIC’s HFI has been associated with child malnutrition and infectious diseases (4). HFI has been associated with discrimination of women in the household suggesting that women are the most vulnerable in this regard (5). FI has also been consistently associated with maternal anemia, domestic violence and depression in women, and poor early childhood development (4)

With the global health focus on the aftermath of the pandemic, food insecurity has been predicted to become the “sting in the tail of COVID-19” (6), hence carefully and efficiently monitoring household food insecurity, in the context of social distancing, has become a global public health priority. The pandemic has become a strong challenge for achieving the Sustainable Developmental Goals (SDG), especially in Low- and Middle-Income Countries (LMIC) where maternal and under-five malnutrition, morbidity, and mortality are expected to rise due to the major poverty increases and disruption of primary health and related services (7). Hence, global health authorities have advised countries to be alert and prepared for a COVID-19-undernutrition syndemic strongly rooted in social determinants of health inequities (8). In 2005, the World Health Organization (WHO) Commission for Social Determinants of Health (CSDH) was established to understand and act on inequities in health (9). One of the three recommendations of the commission to close the gap in a generation was “to measure and understand the problem and assess the impact of action” on inequities in social determinants of health (10). Given that “closing the gap in a generation” would be a challenge with the current COVID-19 pandemic, counties across the globe need to track household food insecurity (HFI) frequently as it is a crucial social determinant of health. This information is necessary for proper targeting and resource allocations through effective food and nutrition security actions.

HFI has been measured indirectly through indicators such as anthropometry, dietary intake surveys, and household food expenditure. The only direct measure is derived from experience-based HFI scales that capture the experience of a household respondent, cognizant of the food situation in the family, about the lack of access to a healthy and nutritious diet as a result of poverty, social deprivation or situations such as natural or man-made disasters (11). The evolution and validation of experience-based HFI assessment tools has been well documented across world regions. The US Household Food Security Survey Module (US-HFSSM, 1995)(12), Household Food Insecurity Access Scale (HFIAS, 2007)(13) and FAO’s *Escala Latinoamericana y Caribeña de Seguridad Alimentaria* (ELCSA) and derived Food Insecurity Experience Scale (FIES) have been validated across a number of different settings and are now being used extensively to track HFI globally (14). However, these scales have not been validated in self-administered form in LMICs which is a major gap in the context of pandemics or other public health emergencies requiring social distancing measures, such as COVID-19.

The objective of this study was to test the validity of a self-applied culturally adapted 8-item version of ELCSA in a large population-based cohort study of pregnant women in Sri Lanka (15) in the context of the COVID-19 pandemic.

## 2.0 Methods

We conducted cognitive and psychometric validation of the self-administered ELCSA for pregnant women in Sri Lanka (ELCSA-P-SL). The study population included pregnant women in their third trimester (32-36 weeks of gestation) of pregnancy enrolled in the Rajarata Pregnancy cohort in Anuradhapura district, Sri Lanka. The study was carried out in February 2020, just before Sri Lanka was affected by the COVID-19 lockdown.

### 2.1 The context

Sri Lanka became an upper middle income country in 2019 with a Gross National Income (GNI) per capita of USD 4060 (16). The average life expectancy at birth for Sri Lankans is 75.5 years (17). The literacy rate of males and females are 93.6% and 91.7% respectively (18). More than 90% of the population has access to safe drinking water and sanitation (18). The prevalence of poverty (i.e., the percentage of the population below the national poverty line) is reported as 4.1% (19). The food security status of the country is reported in a global survey in 2016 indicating that 7.1% and 17.7% of the population is affected by severe and moderate to severe HFI respectively (20). An ongoing prospective cohort study in Sri Lanka provided a unique opportunity to validate a self-reported measure of HFI due to its relatively high maternal literacy rates, universal maternal and child health service coverage and focus in equity for women.

### 2.2 Cognitive validation

The adult 8-items of English version of ELCSA was culturally adapted to Sri Lanka’s context and translated into Sinhala and Tamil following a modified version of Sumathipala and Murray’s method (21). Cognitive validation was performed through expert opinion with five local experts with different experiences in the fields of nutrition, scale development methodology, maternal health, social determinants of health, and public health. Target group interviews were held with ten pregnant women using the methods suggested by Bowden et al for cognitive validation (22). The intended meanings of the eight items of the original questionnaire was compared with the understanding of the translated versions of each of the items. We especially focused on ensuring that the original meaning was preserved; and that the wording was clear, simple and self-explanatory to facilitate its’ self-administration.

### 2.3 Psychometric validation

The cognitively validated ELCSA-P-SL was self-administered by a consecutive sample of 269 pregnant women attending the Rajarata Pregnancy Cohort (RaPCo) special clinics in Anuradhapura district, Sri Lanka, in February 2020 (15). Data were collected from 21 out of 22 public health administrative areas (Medical Officer of Health areas) in the district. Pregnant women filled the questionnaire themselves. Participants psychological distress was simultaneously assessed using the validated Sinhala and Tamil versions of the General Health Questionnaire 12 (GHQ 12)(23) in order to assess the concurrent validity of the new HFI scale. We assessed internal consistency, psychometric properties and fit using a one parameter logistic model (Rasch analysis) using STATA version 14. Item severity scores and infit statistics were calculated using WINSTEP software version 4.3.4.

## 3.0 Results

### 3.1 Changes made in Cognitive validation

Expert and mothers agreed that the questions were clear to them and that the intended meaning of each item was preserved. Based on consensus from the experts, the introduction of the questionnaire was adjusted to include lay language which can be easily understood by rural women. It was clearly mentioned that lack of access to food was being asked in the context of lack of socio-economic resources; and that it did not refer to food restrictions due to dieting, loss of appetite, or pregnancy related symptoms such as nausea, vomiting or heart burn. To maintain the respondents, focus on the time period the phrases “during the past three months” was added at the beginning of each question. To ensure that the exact intended reasons of food insecurity were taken into account, the phrases such as “due to the above mentioned reasons” or “due to less availability of food in your house” were used in each and every item. Using these phrases repeatedly was designed to enable participants to remain focused on accurately answering the ELCSA-P-SL self-administered questions.

### 3.2 Psychometric assessment

#### 3.2.1 Characteristics of the study sample

Of the 328 pregnant women attended the RaPCo special clinics during this period 269 (82%) responded to the questionnaire. The response rate was 82%. The ethnic breakdown and the level of education represented the study population (Table 1).

**Table 1:**
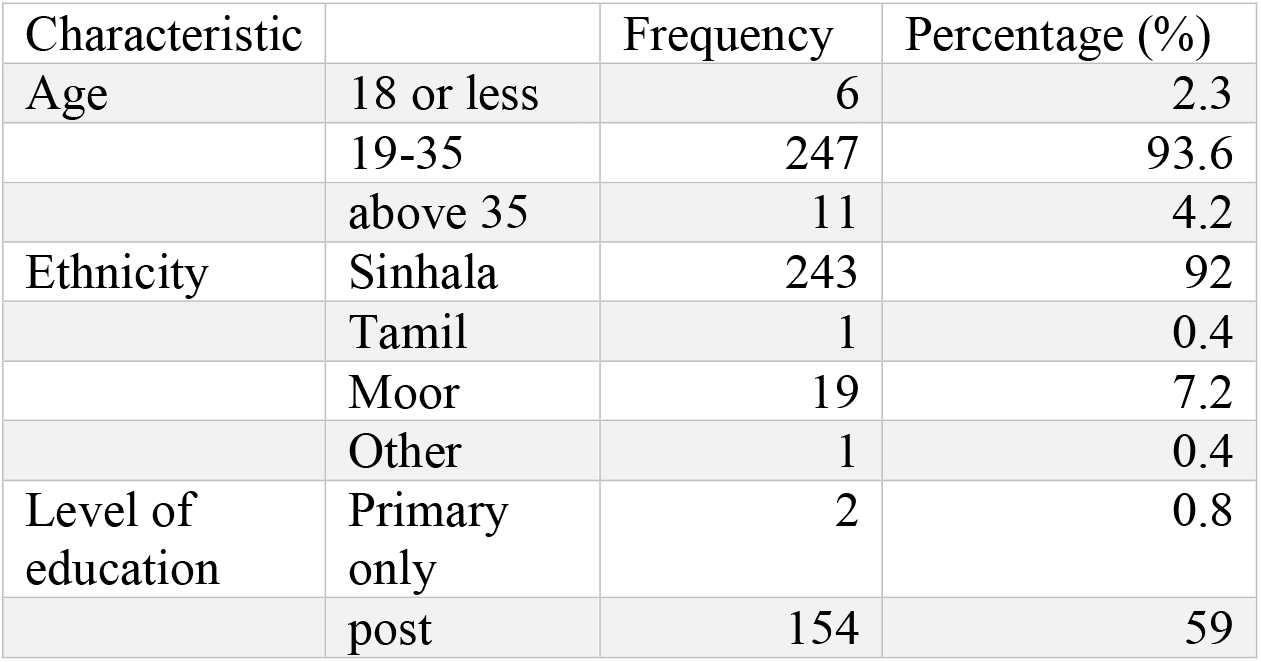
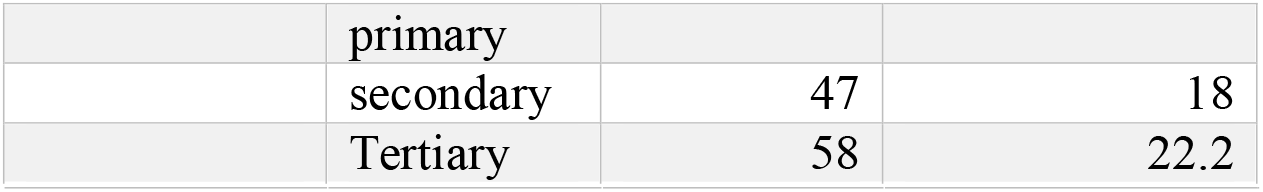
Characteristics of the study population

#### 3.2.2 Frequency distribution of responses

Inability to have a diversity of food due to lack of money or other resources was the item more frequently affirmed. There were no pregnant women in this sample who affirmed item 8; i.e., not eating throughout the day (Table 2). The percentage of missing data ranged from 0.4% to 3.3% in each item. The missing data were replaced by the answer “no” as it did not change the order of severity by doing so and it was predictable based on the pattern of responses in rest of items. As an example a mother who responded “no” to item 7 (experience of hunger for one meal) would invariably not left with hunger during the whole day which indicates a “no” answer for item 8. Further assessment of model fit without replacing the missing data indicated no significant change in severity scores and infit values.

**Table 2:**
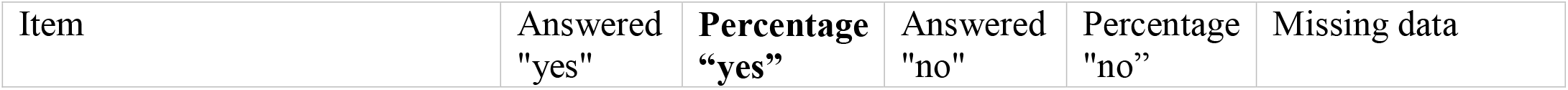
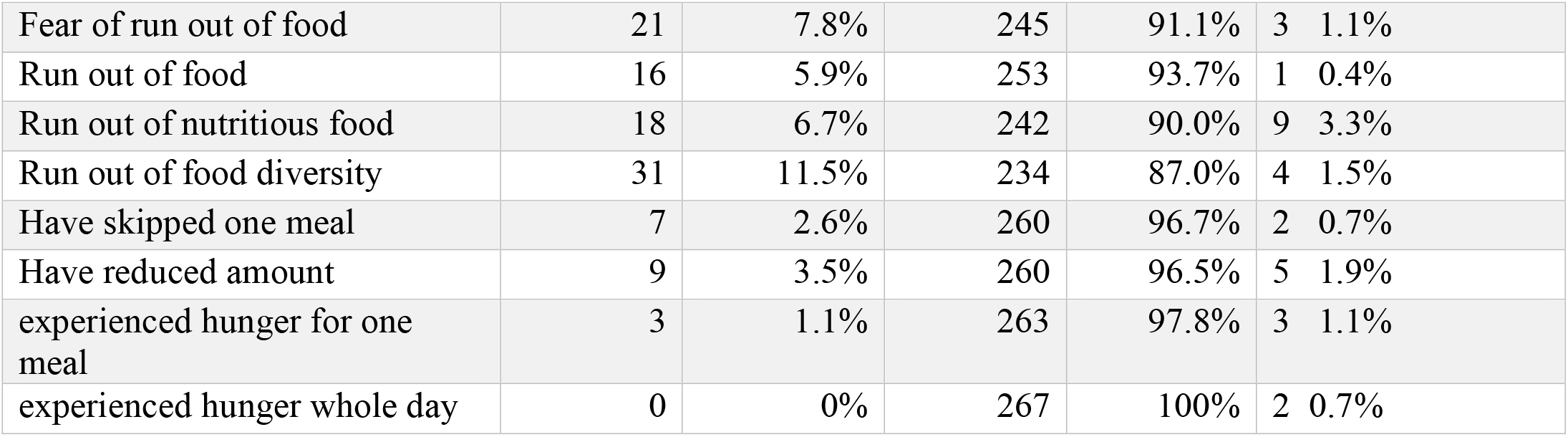
Frequency distribution of responses to the scale

#### 3.2.3 Psychometric properties

The tool was internally consistent (Cronbach’s alpha = 0.79). The one parameter logistic model indicated that food diversity was the first factor to be compromised and that experience of hunger represented the most severe stage of HFI (figure 1). The item infit statistics indicated good model fit (range: 0.85 to 1.07). Item 8 (did not eat for the whole day) was removed from the model fit analysis as no woman affirmed this item. Overall, the item severity scores ranged from -2.15 for diversity food to 4.43 for hunger during the whole day. Concurrent validity between HFI and psychological distress was confirmed (r=0.15, p<0.05).

**Figure 1:**
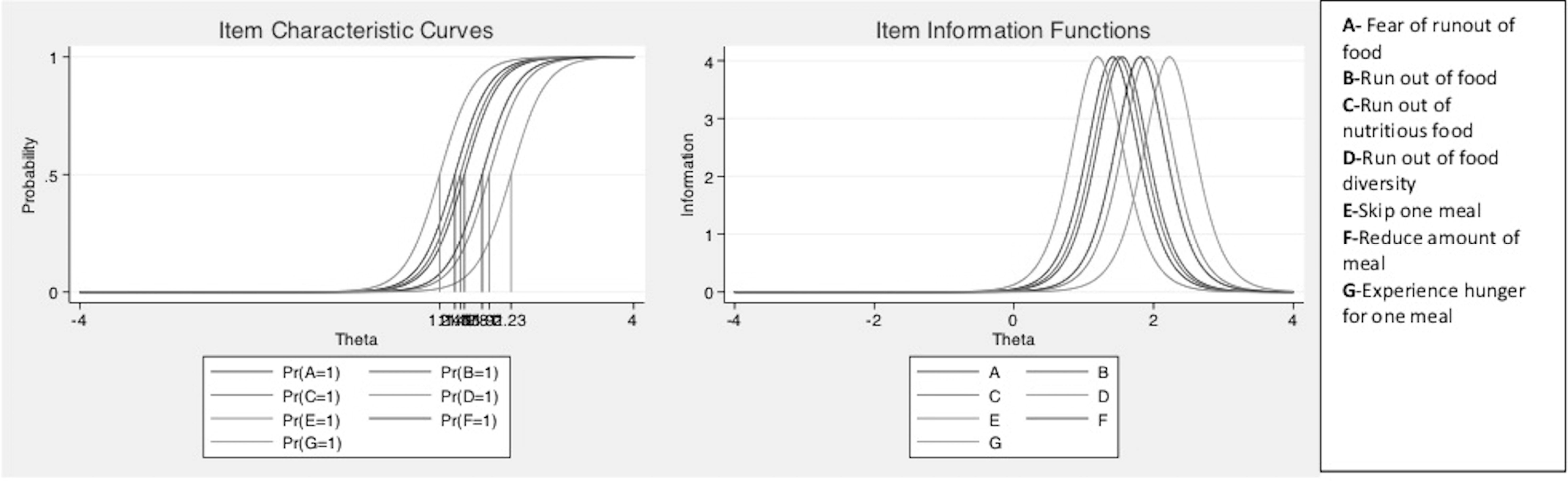
Item characteristic and information curves of ELCSA-P-SL^a^

**Figure 2:**
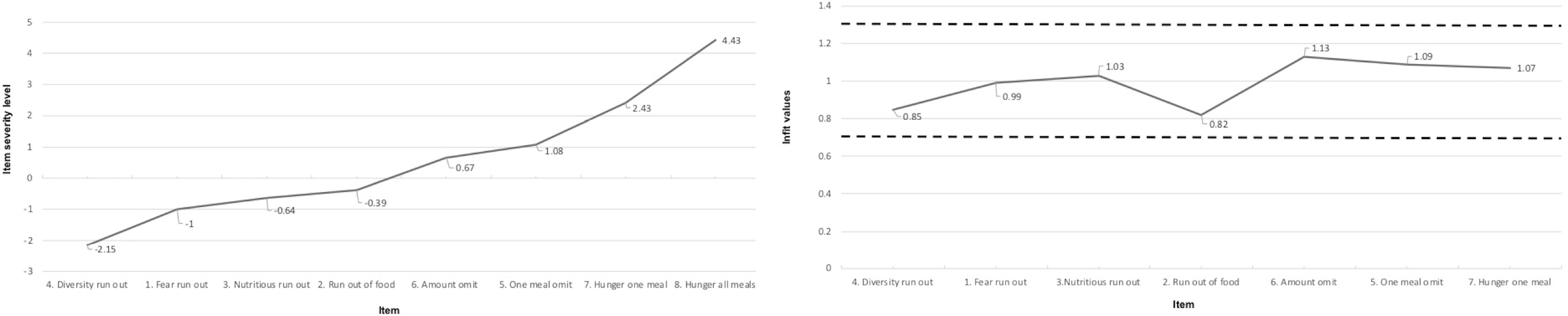
Item severity and fit statistics of ELCSA-P-SL

## 4.0 Discussion

This study reports the validation of the 8-item self-administered ELCSA-P-SL (Appendix 1). The scale had strong cognitive, psychometric, and concurrent validity, hence it can be valuable for fast tracking of HFI in pregnancy in Sri Lanka an similar settings, in the context of social distancing. Although the ELCSA tool had been previously among pregnant adolescents in Brazil (24), as far as we know this is the first time that ELCSA was validated as a self-administered questionnaire among pregnant women in a LMIC country.

The severity scores of ELCSA-P-SL behaved as expected indicating that dietary diversity is sacrificed first and that food intake reduction or hunger is the most severe manifestation of HFI. Poor diet diversity has indeed been identified as being prevalent in Sri Lanka) (25,26). By contrast in other contexts being worried or having fear about running is the item that has been more frequently affirmed (27). Figuring out the reason behind this between-country difference in items’ order require further research.

This study is likely to have minimized social desirability bias as the HFI questionnaire was self-administered (20). Likewise, pregnant women who may have felt shy or uncomfortable reporting food problems in a face-to-face interview might be more open to report them in the self-administered modality.

In Sri Lankan as in many other cultures, mothers are the persons most informed about the food situation in the household. Hence the validity of our findings should not be extrapolated to other possible respondents including fathers or other relatives.

One important advantage of ELCSA is that it not only captures the access to food, but also reflects the underlying pain and anxiety experienced by the participants due to food insecurity and hunger. Indeed, HFI derived from ELCSA-P-SL was significantly correlated with psychological distress which is in agreement with recent findings form HFI phone surveys in Mexico (supplementary file).

Ideally the psychometric validation of ELCSA-P-SL should have been done separately for the Sinhala and Tamil language versions. However, because the sample of Tamil pregnant women was very small (n=20) we were not able to do so. However, we did confirm that removing Tamil participants from the validation analyses led to very similar findings (available upon request from authors) and did not affect any of the conclusions.

The items included in the ELCSA-P-SL scale are not pregnancy specific; thus, in principle this scale could also be used for rapid assessment of HFI in the lactation period or in the general population. Hence further research is needed to assess the external validity of self-administered ELCSA-P-SL to other stages of the woman’s life course or other populations in the context of emergencies that require social distancing measures. This research can help the public health and social protection sectors response to COVID-19 and its aftermath in Sri Lanka and beyond.

## 4.0 Conclusion

The self-administered ELCSA-P-SL is valid and feasible to rapidly track HFI among pregnant women in Sri Lanka.

## Data Availability

Data will be made available by the corresponding author on request.

## Author contributions

RPE, TCA, SBA, GSA, IUJ designed the study. GSA, IUJ, NDW and AUH conducted the research. TCA, SBA and AHF conducted statistical analysis. TCA, NDW and AHF wrote the paper. RPE and SBA edited the manuscript. All authors had the primary responsibility for final content of the data. All authors have read and approved the final manuscript.

## References

1. Laraia BA, Siega-Riz AM, Gundersen C. Household Food Insecurity Is Associated with Self-Reported Pregravid Weight Status, Gestational Weight Gain, and Pregnancy Complications. J Am Diet Assoc [Internet]. 2010;110(5):692–701. Available from: http://dx.doi.org/10.1016/j.jada.2010.02.014

2. Hojaji E, Zavoshy R, Noroozi M, Jahanihashemi H, Ezzedin N, Growth C, et al. Assessment of Household Food Security and its Relationship with Some Pregnancy Complications. J Maz Univ Med Sci. 2015;25(123):87–98.

3. Harris-Fry H, Shrestha N, Costello A, Saville NM. Determinants of intra-household food allocation between adults in South Asia - A systematic review. Int J Equity Health. 2017;16(1):1–21.

4. Pérez-Escamilla R. Food security and the 2015-2030 sustainable development goals: From human to planetary health. Curr Dev Nutr. 2017;1(7):1–8.

5. Panter-Brick C, Eggerman M. Panter-Brick C, Eggerman M. Household responses to food shortages in western Nepal. Hum Organ. 1997;56:190–8. Hum Organ. 1997;56:190–8.

6. Editors. Editorial Food insecurity will be the sting in the tail of COVID-19. Lancet Glob Heal [Internet]. 2018;8(6):e737. Available from: http://dx.doi.org/10.1016/S2214-109X(20)30228-X

7. Roberton T, Carter ED, Chou VB, Stegmuller AR, Jackson BD, Tam Y, et al. Articles Early estimates of the indirect effects of the COVID-19 pandemic on maternal and child mortality in low-income and middle-income countries□: a modelling study. Lancet Glob Heal [Internet]. 2020;(20):1–8. Available from: http://dx.doi.org/10.1016/S2214-109X(20)30229-1

8. Cornia GA, Jolly R, Stewart F. COVID-19 and children, in the North and in the South [Internet]. Innocenti Discussion Papers UNICEF. 2020. Available from: https://www.unicef-irc.org/publications/1087-covid-19-and-children-in-the-north-and-the-south.html

9. Marmot M. Social determinants of health inequalities. Lancet [Internet]. 2005;365(9464):1099–104. Available from: http://www.ncbi.nlm.nih.gov/pubmed/15781105

10. Report of Commissin fo Social Determinants of Health, Organization WH. Closing the gap in a generation: health equity through action on the social determinants of health. Final Report of the Commission on Social Determinants of Health. Geneva: World Health Organization; 2008.

11. Ballard TJ, Kepple AW, Cafiero C. The food insecurity experience scale: development of a global standard for monitoring hunger worldwide. Technical Paper. [Internet]. Rome; 2013. Available from: http://www.fao.org/economic/ess/ess-fs/voices/en/).

12. Food and Agricultural Organization. Declaration of the World Summit on Food Security, World Summit on Food Security. Rome; 2009.

13. Coates J, Swindale, Bilinsky P. Household Food Insecurity Access Scale (HFIAS) for measurement of household food access: indicator guide (v. 3). Washington DC; 2011.

14. Pérez-Escamilla R, Gubert MB RB, Hromi-Fiedler A. Food security measurement and governance: Assessment of the usefulness of diverse food insecurity indicators for policy makers. Glob Food Sec. 2017;14:96–104.

15. Agampodi TC, Wickramasinghe ND, Indika R, Prasanna R, Mudiyanselage P, Jayathilake B, et al. The Rajarata Pregnancy Cohort (RaPCo): study protocol. BMC Pregnancy Childbirth. 2020;20(374):1–13.

16. World Bank. New country classification according to income 2019-2020. 2020.

17. United Nations Population Division. United Nations Population Division. World Population Prospects: 2019 [Internet]. World bank. 2019 [cited 2020 Jan 5]. Available from: https://data.worldbank.org/indicator/sp.dyn.le00.in

18. Department of Census and Statistics Ministry of National Policies and Economic Affairs. Sri Lanka Demographic and Health Survey 2016. Colombo; 2017.

19. Central Bank Sri Lanka. Economic and Social Statistics of Sri Lanka 2019 [Internet]. Available from: https://www.cbsl.gov.lk/sites/default/files/cbslweb_documents/statistics/otherpub/ess_2019_e.pdf

20. Food and Agricultural Organization U. Methods for estimating comparable rates of food insecurity experienced by adults throughout the world. Rome; 2016.

21. Sumathipala A, Murray J. New approach to translating instruments for cross-cultural research: a combined qualitative and quantitative approach for translation and consensus generation. Int J Methods Psychiatr Res. 2000;9(2):87–95.

22. Bowden A et al. Methods for pre-testing and piloting survey questions: illustrations from the KENQOL survey of health - related quality of life. Health Policy Plan. 2002;17(3):322–30.

23. Abeysena H, Jayawardana P, Peiris U, Rodrigo A. Validation of the Sinhala version of the 12-item General Health Questionnaire. J Postgrad Inst Med. 2014;1(1):1–7.

24. Muñoz-Astudillo M, Martínez J, Quintero A, 2010;12(2):173–183. V de la EL y C de SA en gestantes adolescentes [Validating L-A and CL-A food security scale on pregnant adolescents]. RSP (Bogota). Muñoz-Astudillo Mn, Martínez JW, Quintero AR. Validación de la Escala Latinoamericana y Caribeña de Seguridad Alimentaria en gestantes adolescentes [Validating Latin-American and Caribbean Latin-American food security scale on pregnant adolescents]. Rev Salud Publica. 2010;12(2):173–83.

25. Aguayo VM. Complementary feeding practices for infants and young children in South Asia. A review of evidence for action post - 2015. 2017;13(December 2016):1–13.

26. Sirasa F, Mitchell L, Harris N. Dietary diversity and food intake of urban preschool children in North-Western Sri Lanka. 2020;(March):1–17.

27. Perez Escamilla R, Dessalines M, Finnigan M, Pacho H. Household Food Insecurity Is Associated with Childhood Malaria in Rural Haiti 1, 2. 2009;(7).

